# Phenotyping Antidepressant Treatment Response with Deep Learning in Electronic Health Records

**DOI:** 10.1101/2021.08.04.21261512

**Authors:** Yi-han Sheu, Colin Magdamo, Matthew Miller, Sudeshna Das, Deborah Blacker, Jordan W. Smoller

## Abstract

Efficient, accurate phenotyping for antidepressant treatment response in electronic health records (EHRs) could facilitate precision psychiatry applications but remains a challenge. Increasingly, artificial intelligence methods using “deep learning” applied to clinical data have shown promise in complex classification problems. Here, we systematically evaluate the performance of eight deep-learning-based natural language processing models in classifying response to antidepressants in a large real-world healthcare setting. We obtained data spanning 1990-2018 for adults with depression and a co-occurring antidepressant prescription from the EHR data warehouse of the Mass General Brigham healthcare system (n=111,572). Clinical notes were collected for the following time windows after antidepressant initiation: (1) 2 days to 4 weeks, (2) 4–12 weeks, and (3) 12–26 weeks. A stratified random sample of these note sets (total 4,299 across time periods) were manually reviewed to classify response status as “improved” or “no evidence of improvement” in depression symptoms. All models performed well, with areas under the receiver operator curve (AUROC) of at least 0.80. Positive predictive values (PPVs) ranged from 0.72 – 0.91. In general, models incorporating more information-dense and longer text sequences performed better than others. The best performing model (Longformer-large with sliding window) had an AUROC = 0.88 and PPV = 0.84 at a specificity of 0.88. Our results indicate that deep learning methods applied to EHR data can accurately classify antidepressant response in a real-world healthcare setting. Automated treatment response classification may facilitate a range of research and clinical decision support applications.

## INTRODUCTION

Depression is a prevalent psychiatric disorder associated with substantial disability and healthcare costs.^1^ At present, treatment optimization is often a protracted trial-and-error process. The widespread availability of real-world healthcare data in the form of electronic health records (EHRs) offers a promising resource for large-scale research to characterize predictors of treatment response. However, accurately characterizing antidepressant response from EHR data has been a significant challenge. Manual extraction of response data from large health system cohorts is prohibitively costly, time-consuming, and difficult to scale. Therefore, development of an effective method for automated classification of treatment could be highly beneficial.

A major obstacle has been the fact that structured EHR data (e.g., medications, problem lists, ICD billing codes) do not readily provide information relevant to treatment response. For example, Perlis and colleagues (2012) found that the use of billing codes performed only slightly better than chance at classifying antidepressant-related remission of depression (AUROC∼0.55).^2^ However, EHRs also contain a much larger corpus of data in the form of unstructured clinical notes.^3–5^ Efforts to leverage such data have often applied natural language processing (NLP) techniques, which start with the specification of a set of terms associated to the phenotype of interest.^3–5^

Adding NLP on unstructured EHR data to models based on structured EHR data have been shown to enhance antidepressant response classification.^2^ However, the optimal method for incorporating NLP in this setting is unclear. In contrast to conditions that may be well-captured by a specific set of medical terms, text related to depression and treatment response may involve a highly heterogeneous and time-varying sets of terms (including non-specific descriptions of symptoms). In addition, while some response-related text could be encoded by short statements (e.g., “depression: doing well”), it may also depend on extraction of information from longer narrative paragraphs, requiring subtle syntactical and semantical analysis. Classical NLP methods, which typically involve modeling with a “bag of words” (e.g., the presence or counts of a set of words, without consideration of temporal order) fail to account for contextual information.

Recently, deep-learning based NLP models have achieved paradigm-shifting results in general NLP tasks.^6,7^ These models make use of text information beyond seeking a fixed set of terms. Instead, each word in the text is pre-injected with contextual and syntactical information, and the model automatically learns which ones are related to the prediction output. These methods can also accommodate lexical derivatives and typographical errors, without the need for manual curation.

Here, we compare a total of eight deep-learning based NLP modeling approaches to classify antidepressant response during clinically relevant time windows after the first antidepressant treatment was given. Because the vast majority of antidepressant treatments are initiated by primary care clinicians rather than psychiatrists,^8^ we focus on patients started on medication by non-psychiatrists. We show that, despite some differences between models, the deep-learning based NLP models overcome prior limitations that have constrained the classification of antidepressant response outcomes.

For the reader’s convenience, we provide a glossary of technical terms that are either related to deep learning or defined specifically for this work in Table 1.

**Table 1.**
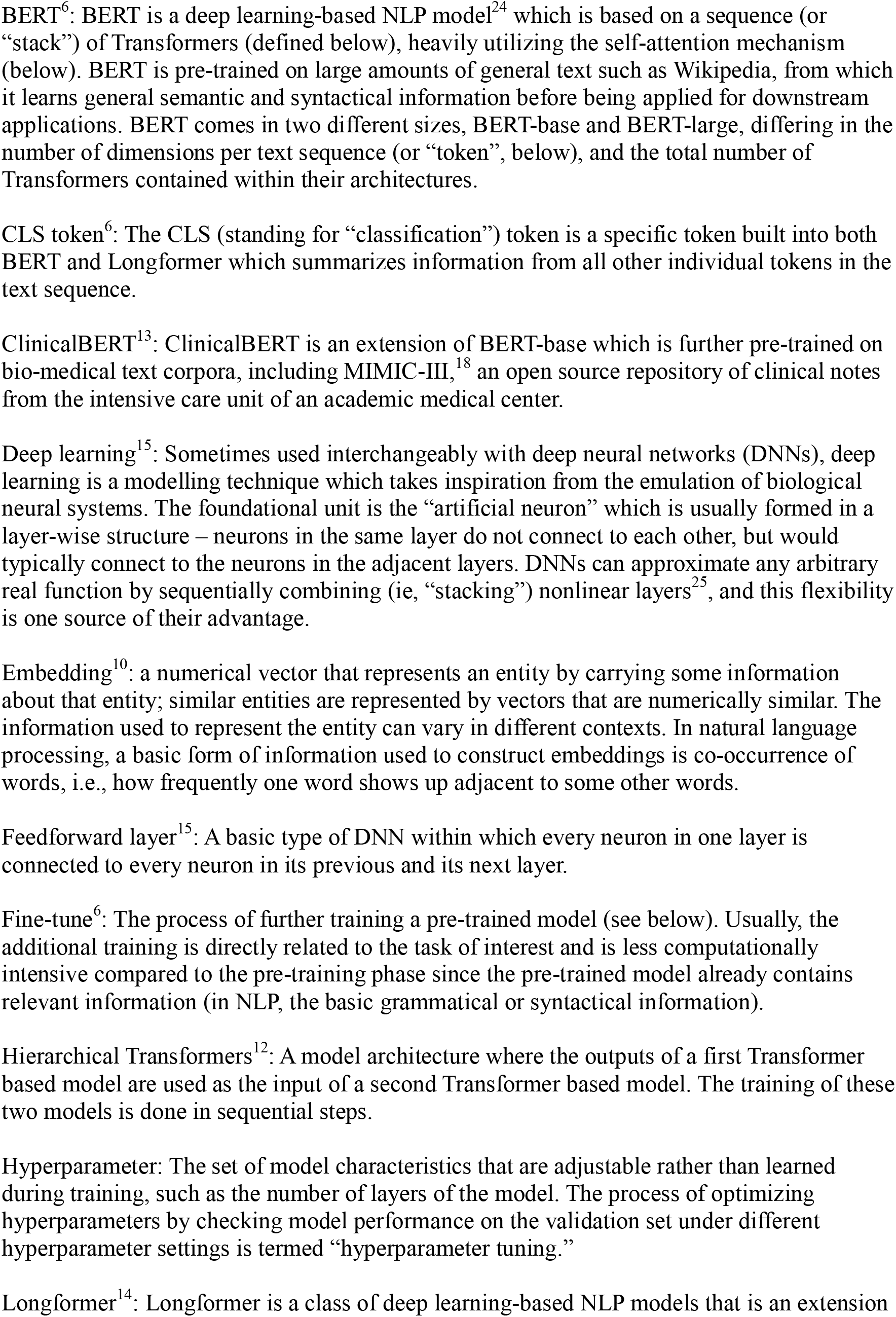
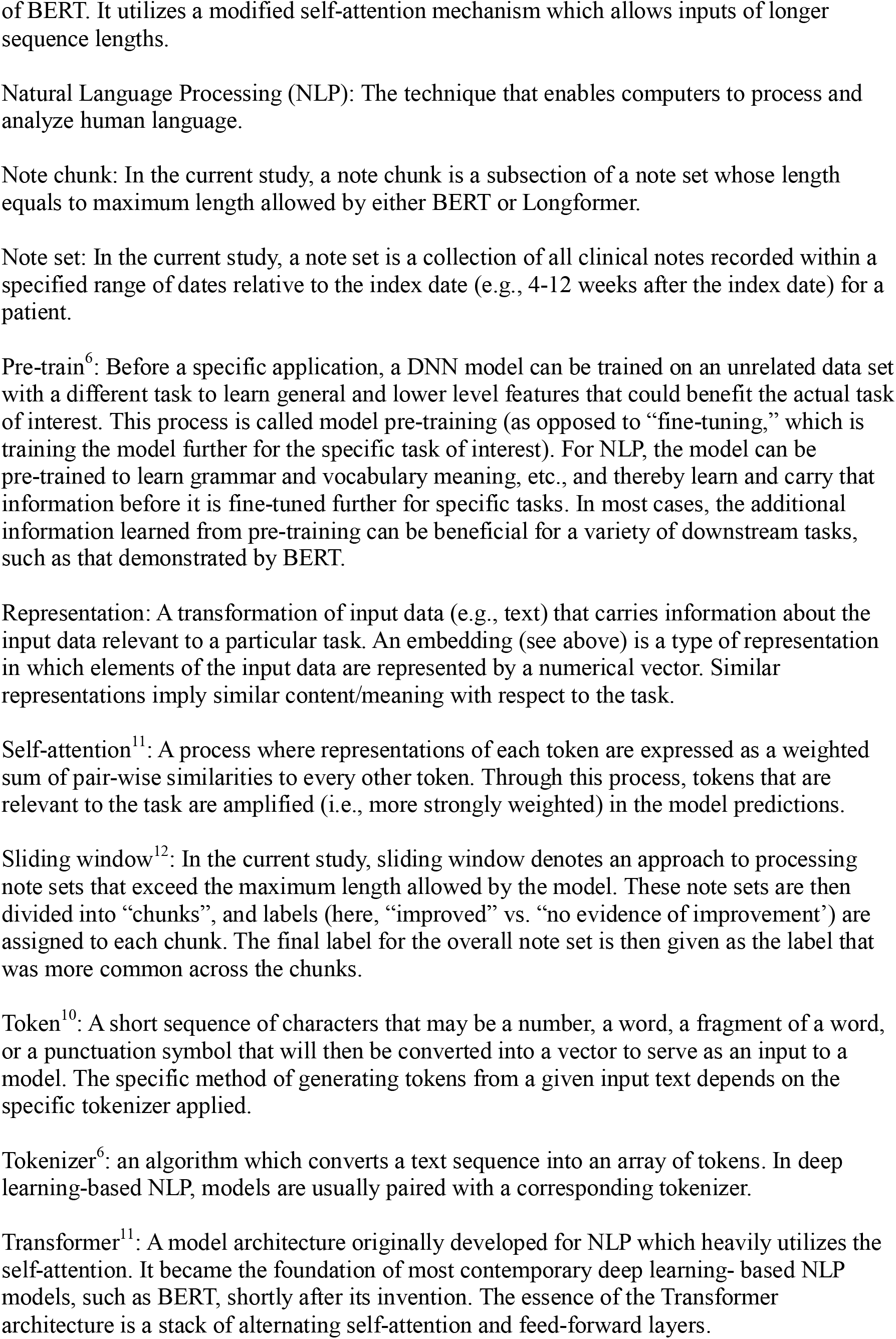
Glossary of terms defined, or related to deep learning appeared in this work

## METHODS

### Institutional Review Board (IRB)

This study was reviewed and approved by institutional review board of the Mass General Brigham (MGB) Healthcare System.

### Data Source

The study data were extracted from unstructured (i.e., free text) clinical note data available in the MGB Research Patient Data Registry (RPDR).^9^ The RPDR is a centralized data warehouse that gathers clinical information from hospitals in the MGB system and currently includes more than 7 million patients with over 3 billion records seen across ten hospitals, including two major Harvard teaching hospitals, Massachusetts General Hospital and Brigham and Women’s Hospital.

Clinical notes were obtained for patients aged 18 or older who had at least one visit with an International Classification of Diseases-Clinical Modification (ICD-CM) code for a depressive disorder (ICD-9-CM: 296.20-6, 296.30-6, and 311; ICD-10-CM: F32.1-9, F32.81, F32.89, F32.8, F33.0-3, and F33.8-9) and at which an antidepressant prescription was initiated during the span of 1990-2018. The notes included office visits, admission notes, progress notes, discharge notes, and correspondence from all medical specialties in the MGB system. The clinical notes come in many varieties and can contain a wide range of information such as the patient’s chief complaint; history of present illness; the physician’s examination and observations; the physician’s assessment and treatment plans, which may or may not include a list of active problems; current and past medications with or without doses; and patient-reported outcome measures, in the form of questionnaires that were converted to text and merged with other sources of clinical notes.

### Data sampling and labeling for Classification of Treatment Response

For labeling and modeling of treatment response classification, we obtained prescription information from the RPDR and identified the first date of prescription (index date) of an antidepressant for each patient. We excluded patients who started more than one antidepressant on the index date because those patients were likely to have been prescribed antidepressants prior to their entry to RPDR. The available notes for each patient were then collected in sets defined by three time windows after the index date: (1) 2 days to 4 weeks, (2) 4–12 weeks, and (3) 12–26 weeks. Notes in each time window for each patient were concatenated as a “note set,” which we defined as the unit of classification (i.e., for human expert labeling and model classification). One of the authors (YHS), a psychiatrist, then manually labeled 628 randomly sampled note sets from time window (1), 3,091 note sets from time window (2), and 580 note sets from time window (3) (total 4,299 note sets). The labeled data were randomly split into a training set of 3,799 note sets, a hold-out validation set of 200 note sets, and a fixed hold-out test set of 300 note sets for which we report final model performance metrics (Figure 1). Note that since available notes from all sources (i.e., not confined to mental health specialty care) were included in the note sets, information regarding depression is typically very sparse.

**Figure 1.**
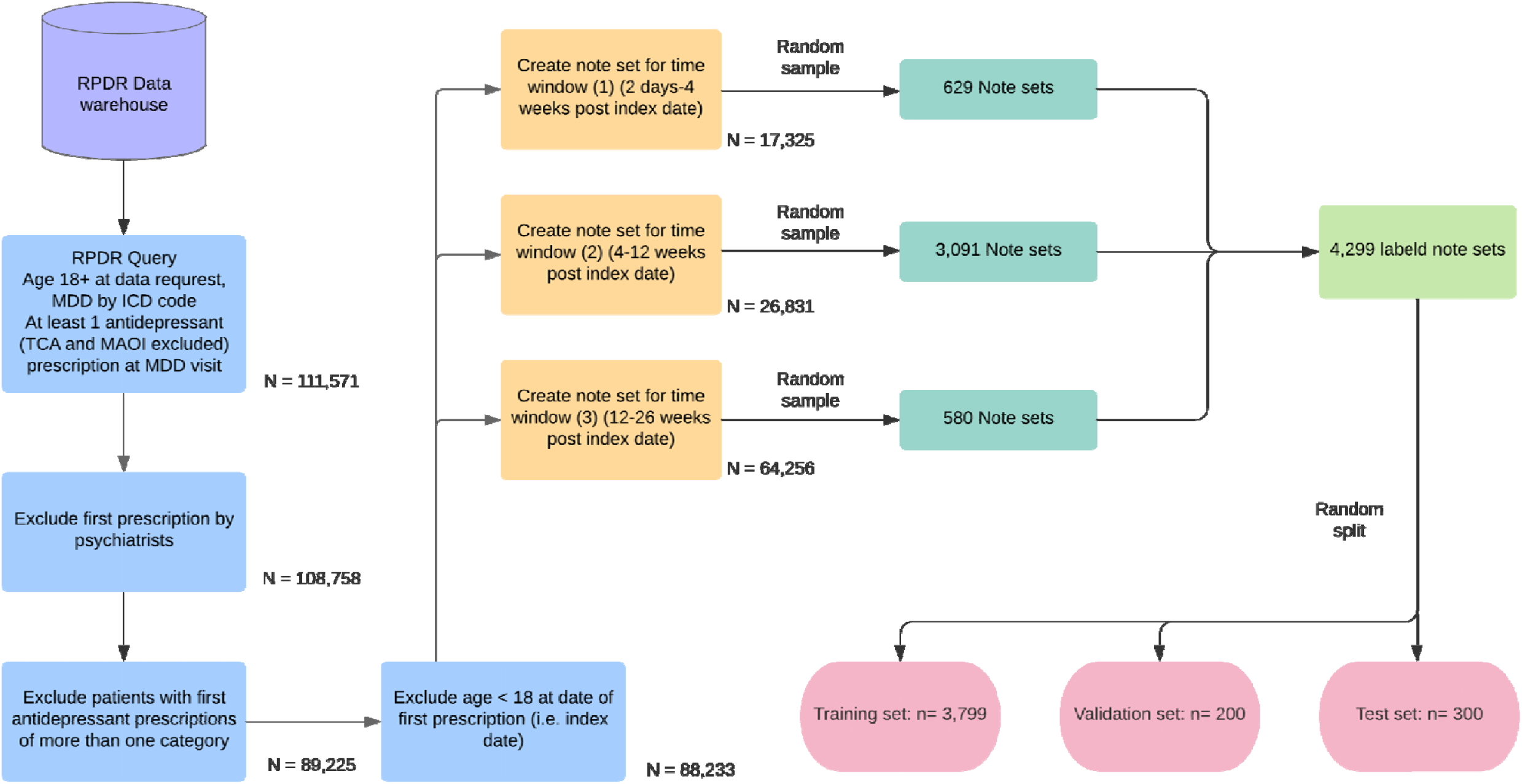
Flow diagram for data retrieval and sampling/construction process of the note sets.

We defined two classes for our classification labels as follows: class (1), Improved: there was evidence in the note set that the patient responded to treatment based on an assessment of their current status, defined by descriptions of depressive symptoms changing in a positive way (e.g., “depression symptoms improved,” “depression well controlled,” or “on X medication and felt much better”); class (2), No evidence of improvement: there was either information in the note set describing mood status, but no evidence of a positive response (e.g., “patient continues to have low mood after starting X medication six weeks ago”) or apparent depression-related symptoms are recorded in the note set without any mention of change (e.g. “this visit, the patient came in stressed and crying, complaining of persisting sleep problems…”), or information is lacking or insufficient in the note set regarding mood status (e.g., there are visits for other conditions that do not provide any assessment of mood or mental status, or the patient’s condition does not allow proper assessment of mood [e.g., due to change in consciousness]), or in the rare case, if the patient was noted to have gotten worse. During labeling, if there were two or more mentions of depression status in the note set over time, the latest one with any description of mood state was chosen for labeling.

### Text preprocessing

In the clinical notes, all non-alphanumeric characters were removed, except for “.” And “-”, which were used by the models. In addition, identifying information (names, medical record numbers, providers, etc.) was removed.

Before the note sets were used as input to a deep learning model, they were first converted to “tokens,” which are short sequences of characters that may be a number, a word, a fragment of a word, or a punctuation symbol (See Glossary for further definition of deep learning terms). Each token was then converted to an “embedding”--that is, a numeric vector that carries prior information about the token.^10^ A simple example of an embedding would be information on how often a given word (e.g. “apple”) co-occurs with other specific words (e.g. “fruit”, “banana”, etc.). Tokens that carry similar information are converted to vectors that are similar. The conversion of text to tokens was done by a “tokenizer.”^6^ The specific choice of tokenizer corresponded to the downstream deep learning model applied.

### Models for Classification of Treatment Response

The model classes we utilized in this study are based on “Transformers.”^11^ The Transformer is a model architecture that heavily utilizes the mechanism of “self-attention.” Self-attention measures similarity between each token in the text, and tokens that are similar and relevant to the task are amplified to produce the final model prediction. This approach has proven to be highly effective for sequential data and is the state-of-the-art approach for current deep-learning based NLP models.^6,11^

However, due to their computational burden, Transformer models often pose an upper limit on the sequence length (i.e., the number of tokens) they can intake at once, and the length of the note sets often exceeded these upper limits (eTable 1). To address this, we adopted the following two approaches (based on Pappagari et. al.^12^): (1) the “sliding window” approach, and (2) the “hierarchical Transformers” approach (Figure 2). We train and optimize models using the training and validation sets, and report test set model performance for four models -- BERT-base,^6^ ClincalBERT,^13^ BERT-large,^6^ and Longformer-large^14^ -- under each of the two aforementioned approaches (a total of 8 models).

**Figure 2.**
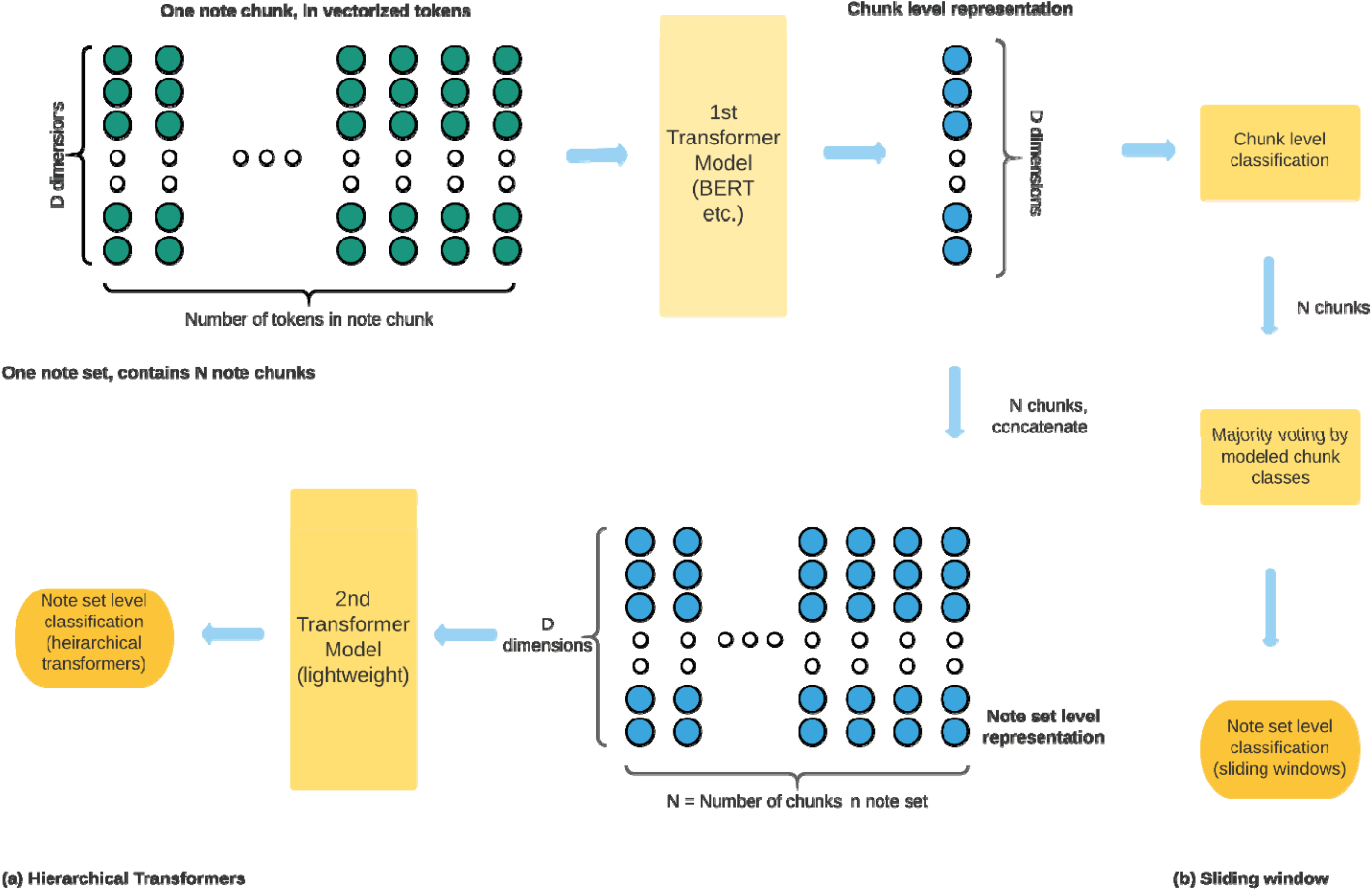
Illustration of information flow for two modeling strategies. Each note set was split into shorter “chunks” for processing. A first level Transformer model was used to derive chunk level representation and classification. (a) The hierarchical Transformers approach (green background) collects all chunk level representations and processes them through a second level lightweight Transformer model to derive note set level classification. (b) The sliding window approach (pink background) collects all chunk level classification labels for a given note set and performs majority voting to derive note set level classification (right hand side). D: number of dimensions used per token. BERT-base and ClinicalBERT: 768, Bert-large and Longformer-large: 1024. N: number of chunks in each note set (which can vary between different note sets).

The BERT model is a large stack of Transformers intertwined with “feedforward layers,”^15^ which are the basic components of deep neural networks whereby all artificial neurons from the previous layer are connected to all of those in the next layer. BERT was “pre-trained” on large scale real world corpora (i.e., Wikipedia and BookCorpus^16,17^), and therefore its model weights already carry semantic and syntactic information about these corpora before BERT is applied to any downstream task. It has demonstrated excellent performance in a variety of NLP tasks^6^ and is still used as a common reference model for NLP tasks. BERT has two versions: BERT-base and BERT-large where BERT-large utilizes a greater number of dimensions representing each token compared to BERT-base (1024 vs 768), allowing more information to be carried in each token. In addition, BERT-large has a larger number of Transformers stacked. ClinicalBERT is a modified version of BERT-base:^13^ in addition to the large corpus of general text, it has been further pre-trained on biomedical texts, including a set of publicly available clinical notes from an intensive care unit, MIMIC-III,^18^ and theoretically carries more healthcare domain-specific information inside its model weights. Longformer is another modification based on BERT, the main appeal being the ability to take in longer sequences compared to BERT, resulting in fewer splits needed when the original text is long. Longformer also comes with a base- and large-format as BERT does. For this study, we adopted the large format (i.e. Longformer-large) which utilizes the same dimensionality per token as BERT-large to examine the effect of ingesting longer sequences and potentially improve performance.

### Information flow and implementation of the classification models

To set up the inputs for each model, the note sets (which can be long) are first split into “chunks,” whose length equals the maximum length each model could intake, with 20% overlapping tokens between a given chunk and its following chunk. Each note chunk inherits the label from its source note set. Two kinds of model outputs were extracted for each note chunk: (1) the modeled probability of the note chunk belonging to the “improved” class, which can be directly converted to a modeled binary label for the chunk, and (2) an embedding” of the note chunk (the output “CLS token”^6^), which is an array of decimal numbers containing information representing the note chunk (related to antidepressant treatment response).

For the sliding window approach, modeled note set labels were assigned using “majority voting” by the modeled note chunk labels in the note set. For the “hierarchical Transformer” approach, a second level light-weight Transformer model (eFigure 1) took in the embeddings of the note chunks produced by the first level model as its input, and outputted modeled note set labels. Each of the eight models was trained (or “fine-tuned”^6^ because some models are pre-trained), and hyperparameters were optimized to maximize note set level accuracy (i.e. the proportion of agreement between the modeled and ground truth labels of the note set) on the training/validation set. The modeling implementations were done using the Python programming language, and deep learning/NLP frameworks PyTorch^19^, Pytorch-lighting^20^, Huggingface Transformers^21^, SimpleRepresentations,^22^ and SimpleTransformers^23^.

### Model performance metrics

For each model, we report the following performance on the hold-out test set: area under the receiver operating characteristic curve (AUROC) and area under the precision-recall curve (AUPRC – PRC denotes the curve where the x-axis is 1-specificty and the y-axis is positive predictive value (PPV)), both of which are threshold agnostic, as well as accuracy, PPV, negative predictive value (NPV), sensitivity, specificity, and F1 score (= 2 * (PPV * sensitivity)/(PPV + sensitivity)), based on a threshold chosen to maximize accuracy. Confidence intervals for the metrics were calculated based on bootstrapping the test set’s predicted probabilities and associated ground truth labels 500 times for each model.

## RESULTS

Demographic characteristics of the sample are shown in Table 2. Point estimates of classification performance metrics of each of the eight models, using either sliding windows or hierarchical Transformers with BERT-base, ClinicalBERT, BERT-large, and Longformer-large, are presented in Table 3. Performance metrics with confidence intervals are provided as eTable 2. ROC curves for all eight models are provided in eFigure 2, and PRC curves are provided in eFigure 3.

**Table 2.**
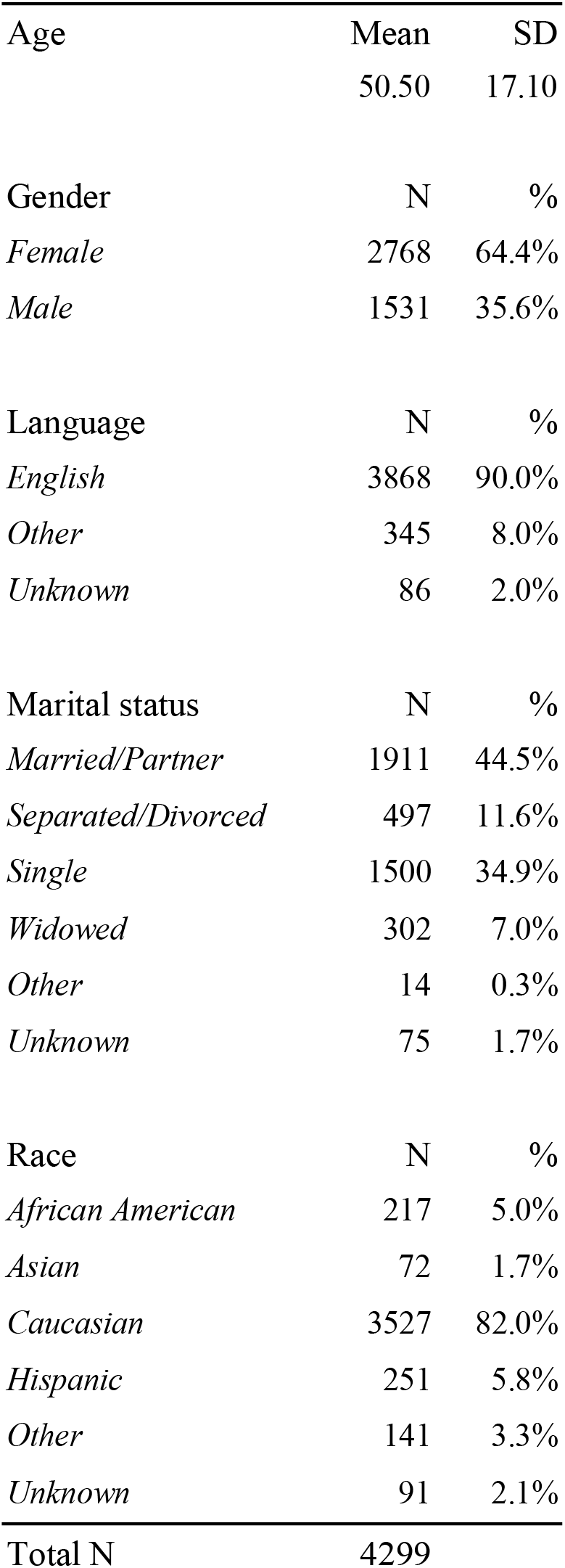
Distribution of key demographics variables among for the patients constituting the note sets used for antidepressant treatment response phenotyping in the current study

|  |  |  |
| --- | --- | --- |
| Age | Mean | SD |
|  | 50.50 | 17.10 |
| Gender | N | % |
| *Female* | 2768 | 64.4% |
| *Male* | 1531 | 35.6% |
| Language | N | % |
| *English* | 3868 | 90.0% |
| *Other* | 345 | 8.0% |
| *Unknown* | 86 | 2.0% |
| Marital status | N | % |
| *Married/Partner* | 1911 | 44.5% |
| *Separated/Divorced* | 497 | 11.6% |
| *Single* | 1500 | 34.9% |
| *Widowed* | 302 | 7.0% |
| *Other* | 14 | 0.3% |
| *Unknown* | 75 | 1.7% |
| Race | N | % |
| *African American* | 217 | 5.0% |
| *Asian* | 72 | 1.7% |
| *Caucasian* | 3527 | 82.0% |
| *Hispanic* | 251 | 5.8% |
| *Other* | 141 | 3.3% |
| *Unknown* | 91 | 2.1% |
| Total N | 4299 |  |

**Table 3.** Classification performance metrics for each model on the hold-out test set (n=300). For threshold-dependent metrics (accuracy, F1, NPV, PPV, sensitivity, specificity), for each model we chose the model probability threshold that maximized the accuracy of label binarization. The highest values for each column were bolded for clarity. Hierarchical: Hierarchical Transformers modeling approach.

| Model | Strategy | Accuracy | AUROC | AUPRC | F1 | NPV | PPV | Sensitivity | Specificity | Threshold |
| --- | --- | --- | --- | --- | --- | --- | --- | --- | --- | --- |
| BERT-base | *sliding window* | 0.75 | 0.81 | 0.78 | 0.72 | 0.77 | 0.72 | 0.73 | 0.76 | 0.512 |
|  | *hierarchical* | 0.78 | 0.84 | 0.80 | 0.73 | 0.76 | 0.82 | 0.66 | 0.88 | 0.567 |
| ClinicalBERT | *sliding window* | 0.74 | 0.80 | 0.75 | 0.70 | 0.75 | 0.74 | 0.68 | 0.80 | 0.551 |
|  | *hierarchical* | 0.76 | 0.82 | 0.79 | 0.69 | 0.73 | 0.83 | 0.60 | 0.90 | 0.499 |
| BERT-large | *sliding window* | 0.77 | 0.84 | 0.83 | 0.76 | 0.81 | 0.72 | **0.79** | 0.75 | 0.453 |
|  | *hierarchical* | 0.82 | 0.86 | 0.81 | **0.78** | 0.78 | 0.89 | 0.68 | 0.93 | 0.554 |
| Longformer -large | *sliding window* | **0.83** | **0.88** | **0.87** | 0.81 | **0.83** | 0.84 | 0.78 | 0.88 | 0.502 |
|  | *hierarchical* | 0.82 | 0.84 | 0.85 | 0.77 | 0.77 | **0.91** | 0.66 | **0.95** | 0.533 |

All eight models performed well, achieving an AUROC greater than 0.80. Overall, the best performing model was Longformer-large with sliding windows: AUROC 0.88, AUPRC 0.87, PPV 0.84, and NPV 0.83 with a sensitivity of 0.78, specificity of 0.88, and F1 score of 0.81 at a decision threshold maximizing accuracy.

We also examined the impact of four key modeling decisions with pair-wise comparisons between contrasting models:

### (1) Effect of pre-training the model on bio-clinical text (BERT-base vs ClinicalBERT)

ClinicalBERT shares the same model structure as BERT-base, but is expected to carry more domain-specific information. Although the differences were small (by point estimates of model metrics), BERT-base counter-intuitively performed better than ClinicalBERT under both sliding windows and hierarchical approaches.

### (2) Effect of larger input vector size per token and larger number of stacked Transformers (BERT-base vs BERT-large)

BERT-base and BERT-large share identical model architecture and pre-training methods but differ in the dimensions used per input token and the number of Transformer encoders stacked. As expected, BERT-large consistently outperformed BERT-base, presumably due to its capacity to carry more information.

### (3) Effect of larger maximum sequence length (BERT-large vs Longformer-large)

BERT-large and Longformer-large perform similarly under the hierarchical approach, while Longformer-large performed better with the sliding window approach. With hierarchical Transformers, both models are concatenations of two Transformer-based models, and the difference is mainly concerned with the arbitrary interruption of information flow during training. BERT-large intakes chunks of shorter lengths with a larger number compared to Longformer, so there are more arbitrary cuts in the original text, but at the same time more total information was retained when the chunk representations enter the second level Transformer, and more interaction can be modeled between the chunks. With sliding window, since this is a simplistic rule that does not consider the effects of factors such as timing, the model which could accommodate longer sequences (i.e., fewer splits of text) would retain and process more information in the Transformer model before the majority vote.

### (4) Examining the effect of sliding window vs hierarchical Transformer

Model performance was generally superior with *hierarchical Transformers*, with the exception of Longformer-large. Except for Longformer-large, the other three model classes benefit from the increased complexity from the second level Transformer. Notably, the number of text chunks under the Longformer setting is far smaller than the other three approaches (median number of chunks = 1, eTable 3).

## DISCUSSION

We used data from a large healthcare system to evaluate the performance of newer deep-learning based NLP methods to classify antidepressant response, a particularly complex phenotype that relies on context-dependent patient and clinician reports without the benefit of laboratory indicators. We systematically compared eight deep-learning NLP modeling approaches that varied in their handling of textual inputs. We found that, despite the sparse nature of relevant information captured in clinical notes, these methods performed well overall, achieving AUROCs uniformly greater than 0.80 and PPVs of 0.72 – 0.91. In general, we find that models that incorporate larger size per token and longer input sequences perform best. For example, the Longformer-Large model using a hierarchical strategy for handling text achieved a PPV of 0.91 at a specificity of 0.95. That is, there is more than a 90% probability that a patient classified as improved with antidepressant treatment would in fact be improved.

Several aspects of these results are noteworthy. This study is the first, to our knowledge, to apply deep-learning-based NLP to classify phenotype antidepressant response in EHRs, and demonstrates that this approach can perform very well. One previous study^2^ attempted to classify remission status (but not improvement) among patients with treatment-resistant depression using clinical notes from outpatient psychiatry practices. In comparison, our approach: (1) primarily included clinical notes from non-specialist practices, where mental health information is more sparse, creating a more challenging modeling task; (2) provided higher PPV and sensitivity at the same level of specificity (e.g., Longformer-large, hierarchical approach: PPV: 91% vs 78%, sensitivity: 66% vs 39%, at 95% specificity); and (3) explicitly classifies improvement (rather than just remission) relative to baseline.

Conventionally, classifying phenotypes based on EHRs has relied on one or more of the following methods: 1) human expert review, which is not readily scalable; 2) term-based NLP, which is effectively an automated word search;^2^ and/or 3) use of surrogate information e.g. ICD codes or other structured EHR data, which may not capture the complex and nuanced components of antidepressant response.^3–5^ In contrast, the approach described here is highly scalable, more flexible than term-based NLP (by not requiring pre-specified terms), and takes advantage of the depth and granularity of clinically-relevant information embedded in narrative notes. Compared to term-based NLP or the use of only structured data, the current approach more closely emulates manual chart review process– i.e., an artificial agent “reads through” the notes in a way analogous to a human expert reviewer, taking in the same body of information. Since the information source is identical, this approach can theoretically match the performance of expert review as modeling architectures continue to rapidly improve.^6,7,11^

In addition to phenotyping for treatment response, this “emulation” approach could be used to facilitate automated EHR phenotyping of a broad range of conditions and may be particularly useful for psychiatric applications, where relevant information is often primarily recorded in free text using variable or inconsistent terms. In particular, key information may be incorporated in narratives about symptom change, documented discussions between clinician and patient, descriptions of life events, and assessment and interpretations by the clinician, all of which can be extracted through the modeling approach described here.

Our results should be interpreted in light of several limitations. First, although our method can capture more nuanced information than pre-specified term-based NLP, we are still limited to information documented in clinical notes that may not fully capture relevant features. Second, like any supervised learning approach, our method still requires some degree of manual chart review to derive labels for model training. Third, clinical documentation may vary between systems, so the specific model derived here may not fully generalize to other health systems. If that is true, implementation of our approach would require model re-training at other sites. Lastly, the models were trained supervised by chart reviews, and therefore ultimately constrained by information available in the EHR.

In summary, we applied a set of deep-learning based approaches to phenotype antidepressant treatment response using real-world clinical notes and demonstrate that, despite challenges from sparse information and noisy formats, these approaches achieve high levels of accuracy. With a specificity of 95%, we can achieve a positive predictive value for antidepressant response classification exceeding 90%. With further refinement, this method could enable high-throughput, semi-automated treatment phenotyping that would unlock a range of research and clinical applications.

## Data Availability

The electronic health records data used in this study are not available to the public or per request for privacy protection.

## ACKNOWLEDGEMENTS

The authors thank the MGB RPDR team for providing the EHR data in an analyzable format.

---

**eTable 1.** Percentiles of lengths per note set by tokens. The BERT family (BERT-base, BERT-large, and ClinicalBERT) shares a common way of building tokens and therefore have the same number of tokens per note. Longformer uses a different way to build the tokens.

| Percentile | Token length per note set, BERT | Token length per note set, Longformer |
| --- | --- | --- |
| 0.01 | 138 | 126 |
| 0.1 | 559 | 496 |
| 0.25 | 1065 | 946 |
| 0.5 | 2514 | 2216 |
| 0.75 | 6536 | 5819 |
| 0.9 | 14576 | 12871 |
| 0.99 | 52365 | 46865 |

**eTable 2.**
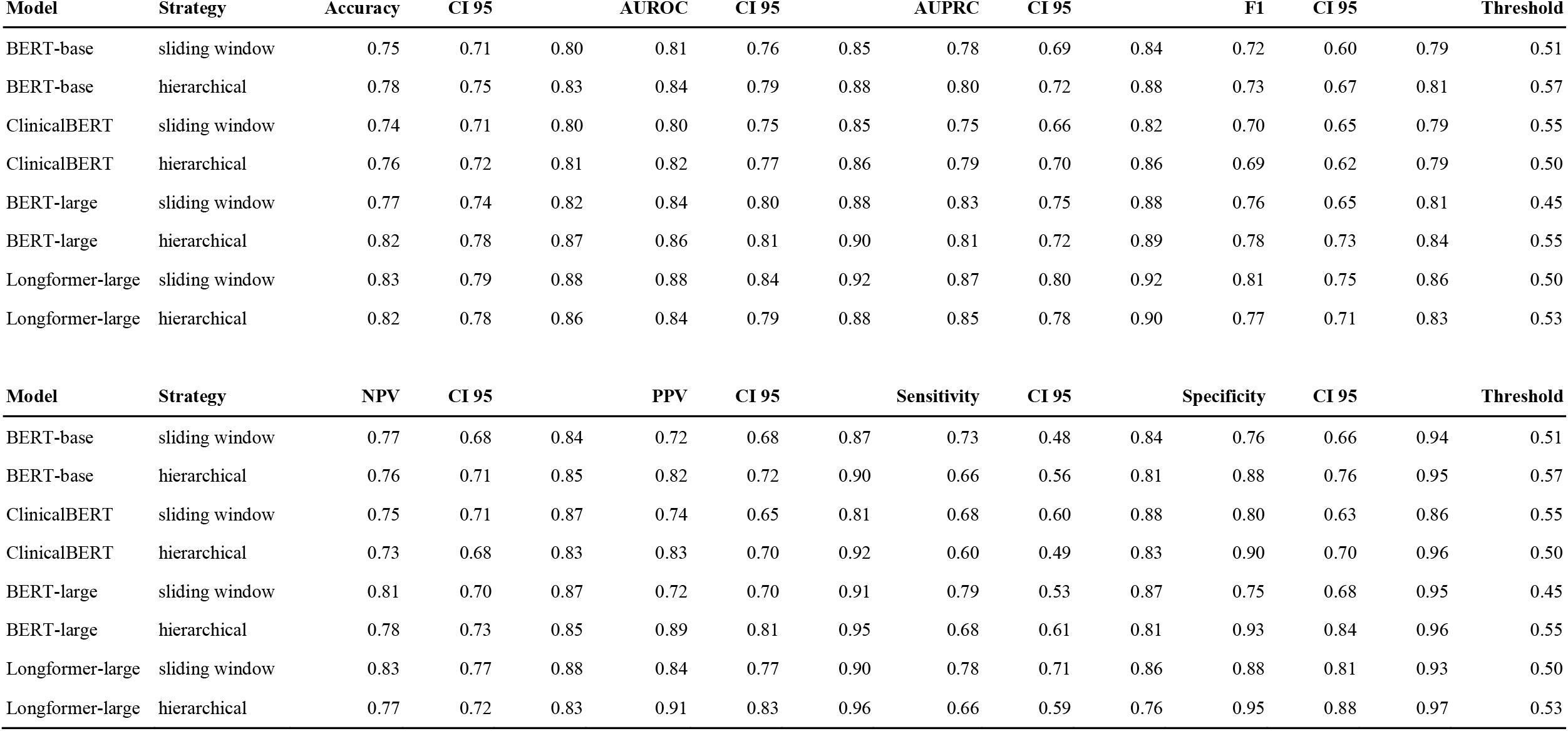
Model performance result metrics with 95% bootstrapping confidence interval.

**eTable 3.** Percentiles of chunks per note set. The BERT family (BERT-base, BERT-large, and ClinicalBERT) shares a common way of building tokens. They also have a smaller limit of sequence length (512) and therefore resulted in more chunks per note set compared to Longformer-large.

| Percentile | Number of chunks per note set, BERT | Number of chunks per note set, Longformer |
| --- | --- | --- |
| 0.01 | 1 | 1 |
| 0.1 | 2 | 1 |
| 0.25 | 3 | 1 |
| 0.5 | 7 | 1 |
| 0.75 | 17 | 2 |
| 0.9 | 37 | 4 |
| 0.99 | 132 | 15 |

## eFigure legends

**eFigure 1.**
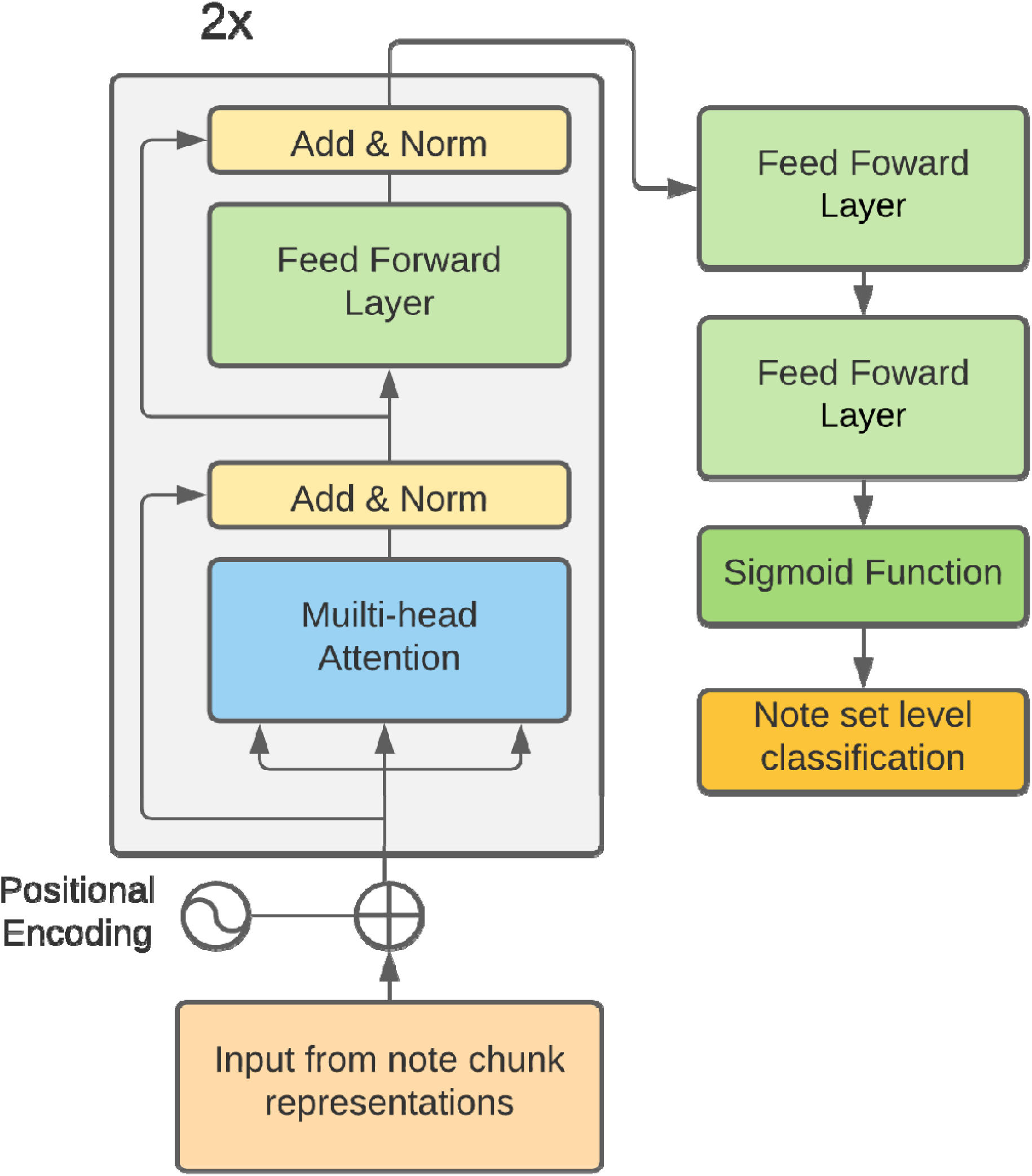
Model architecture of the “lightweight” Transformer applied in the hierarchical Transformer approach. Positional encoding: adding positional marker into the input sequence which enables the model to identify the position of each input token. Norm: normalization to range between 0 and 1. The Transformer architecture (gray box) was described in Vaswani et. al. 2017.^11^

**eFigure 2.**
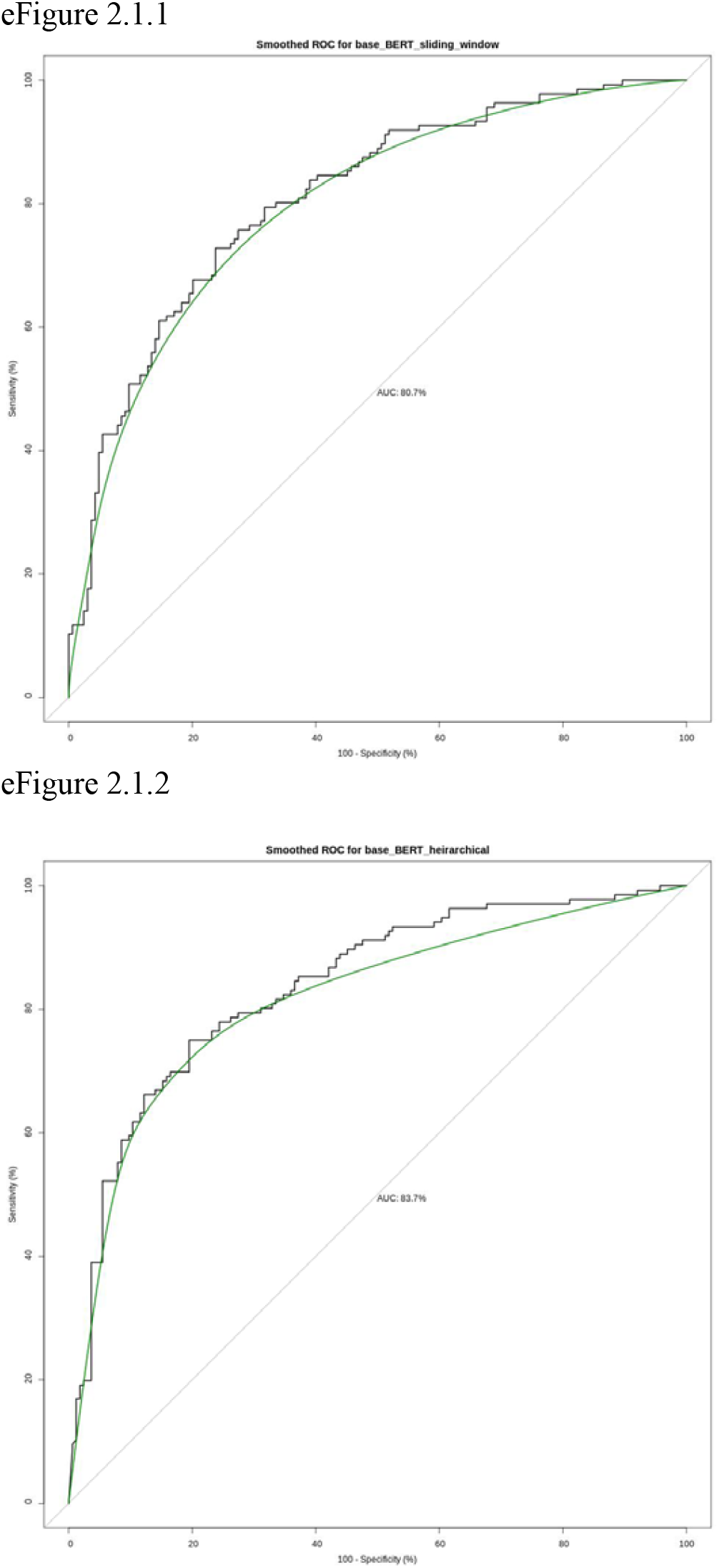
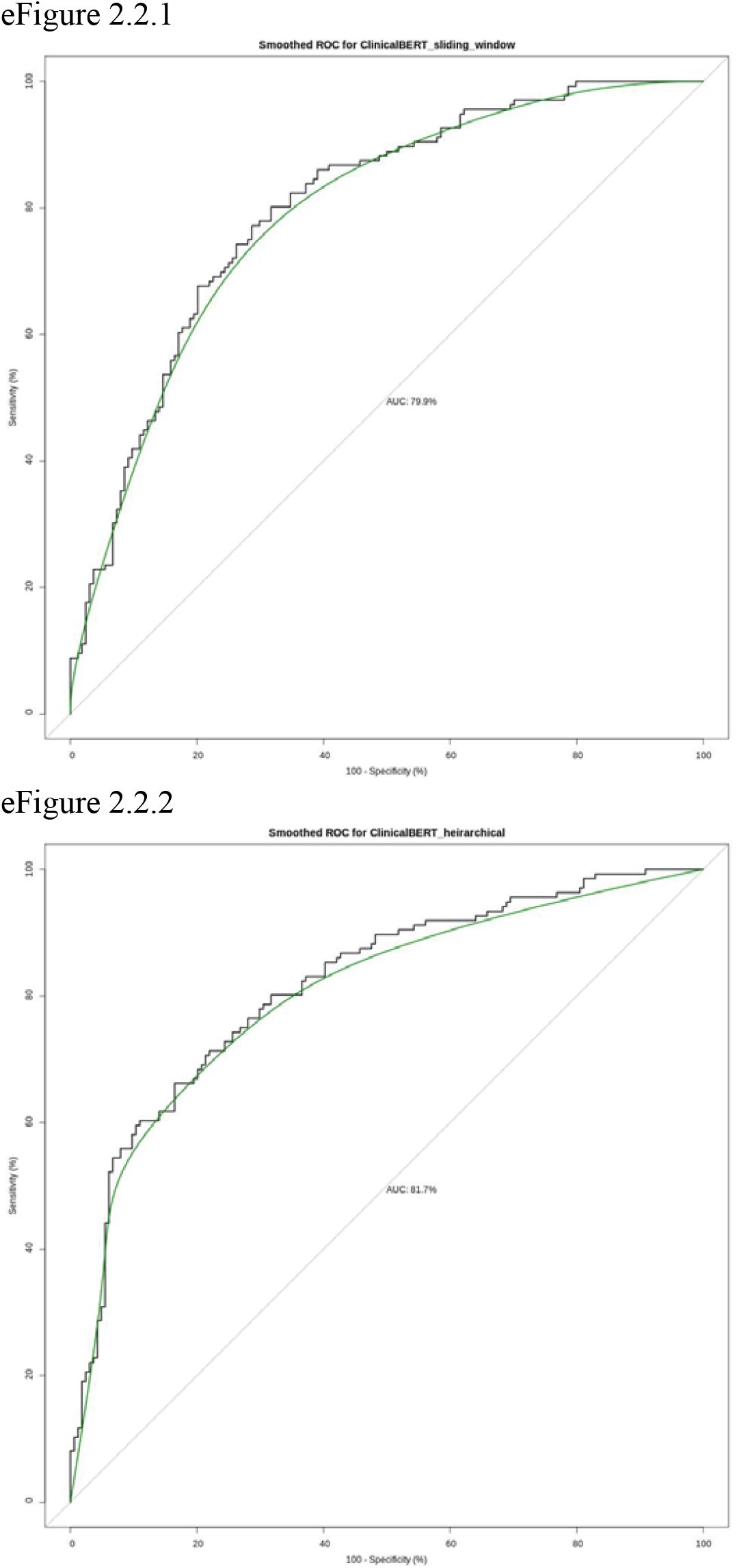
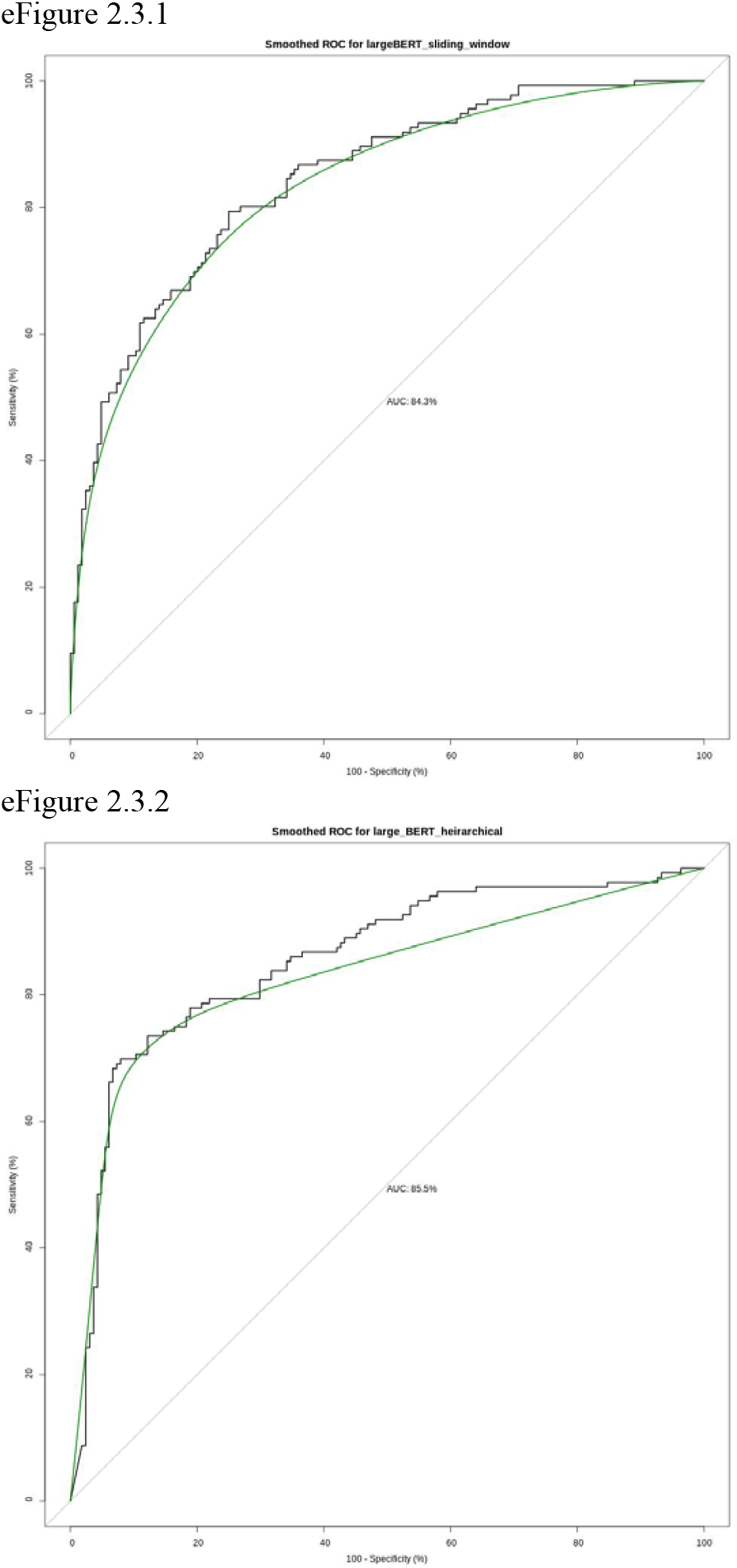
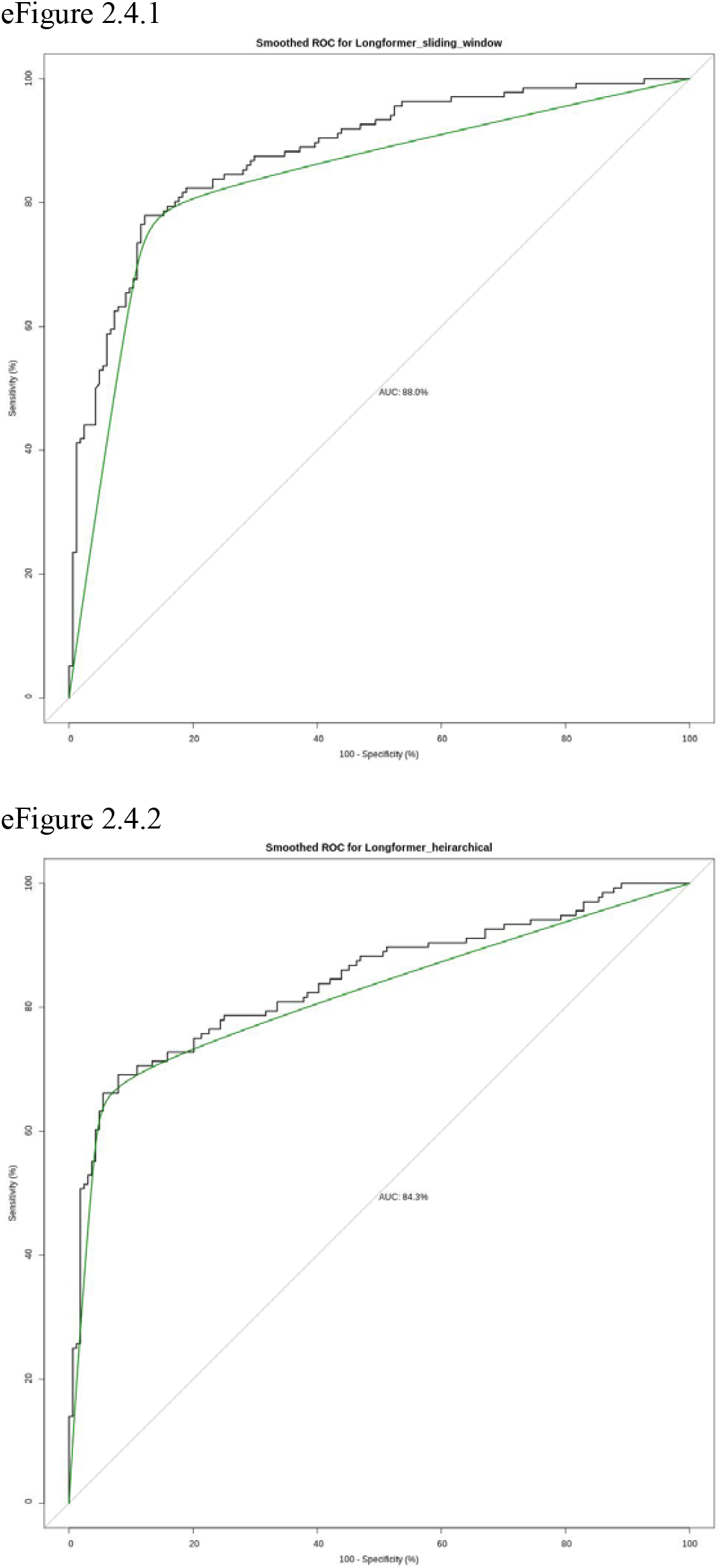
ROC plots for the eight models. e2.1.1 – BERT-base, sliding window; e2.1.2 – BERT-base, hierarchical Transformers; e2.2.1 – ClinicalBERT, sliding window; e2.2.2 – ClinicalBERT, hierarchical Transformers; e2.3.1 – BERT-large, sliding window; e2.3.2 – BERT-large, hierarchical Transformers; e2.4.1 – Longformer, sliding window; e2.4.2 – Longformer, hierarchical Transformers.

**eFigure 3.**
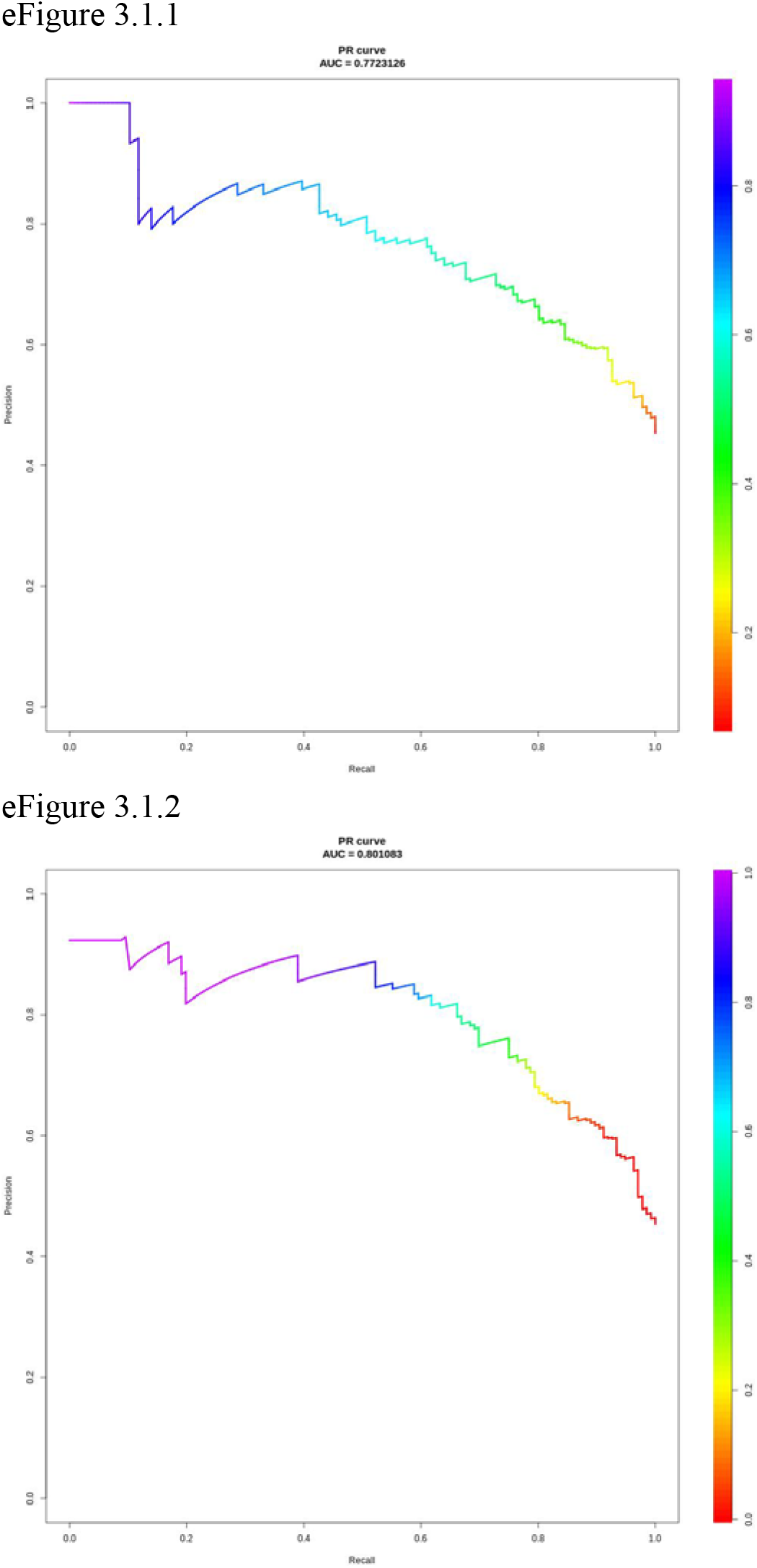
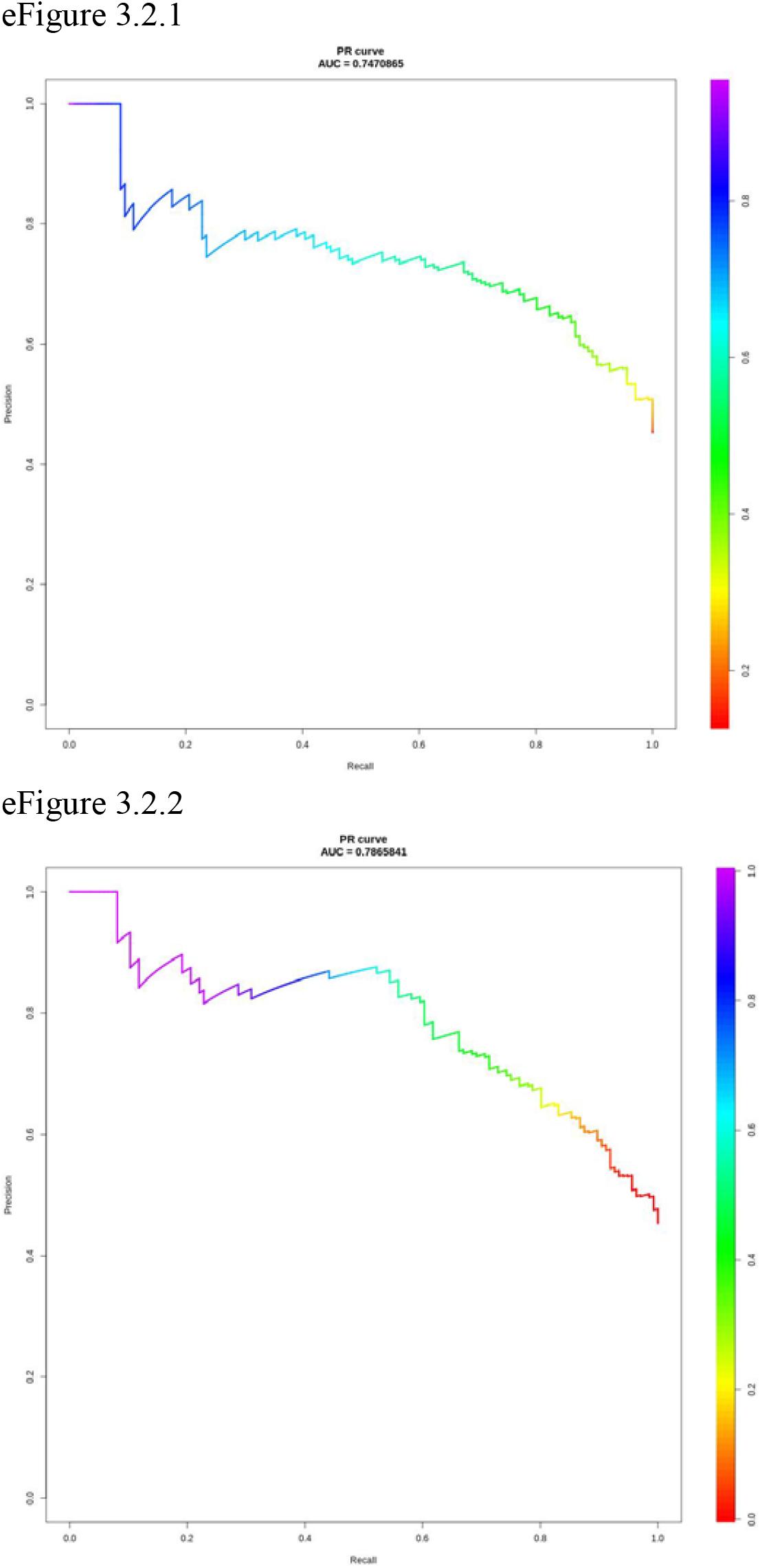
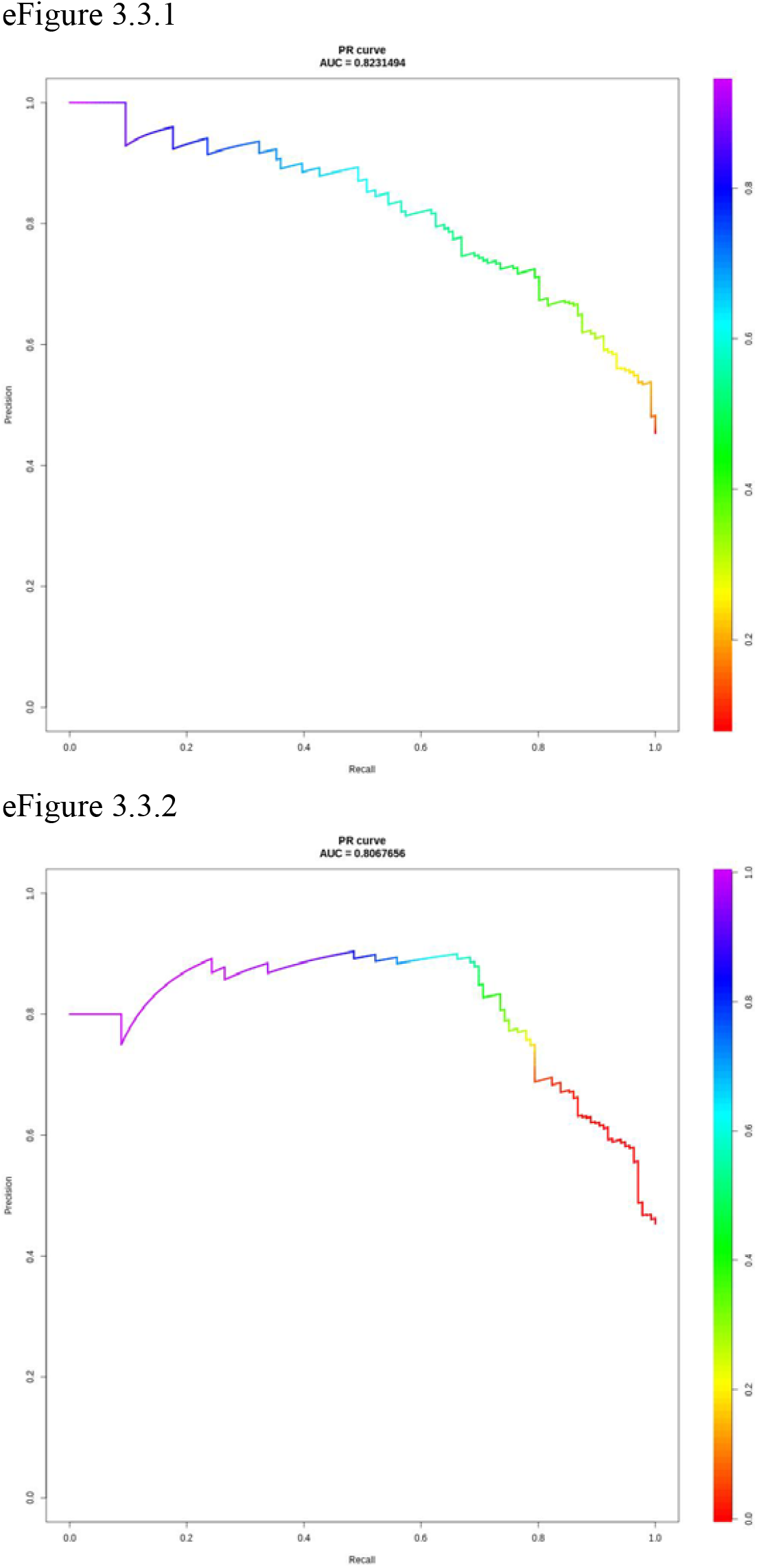
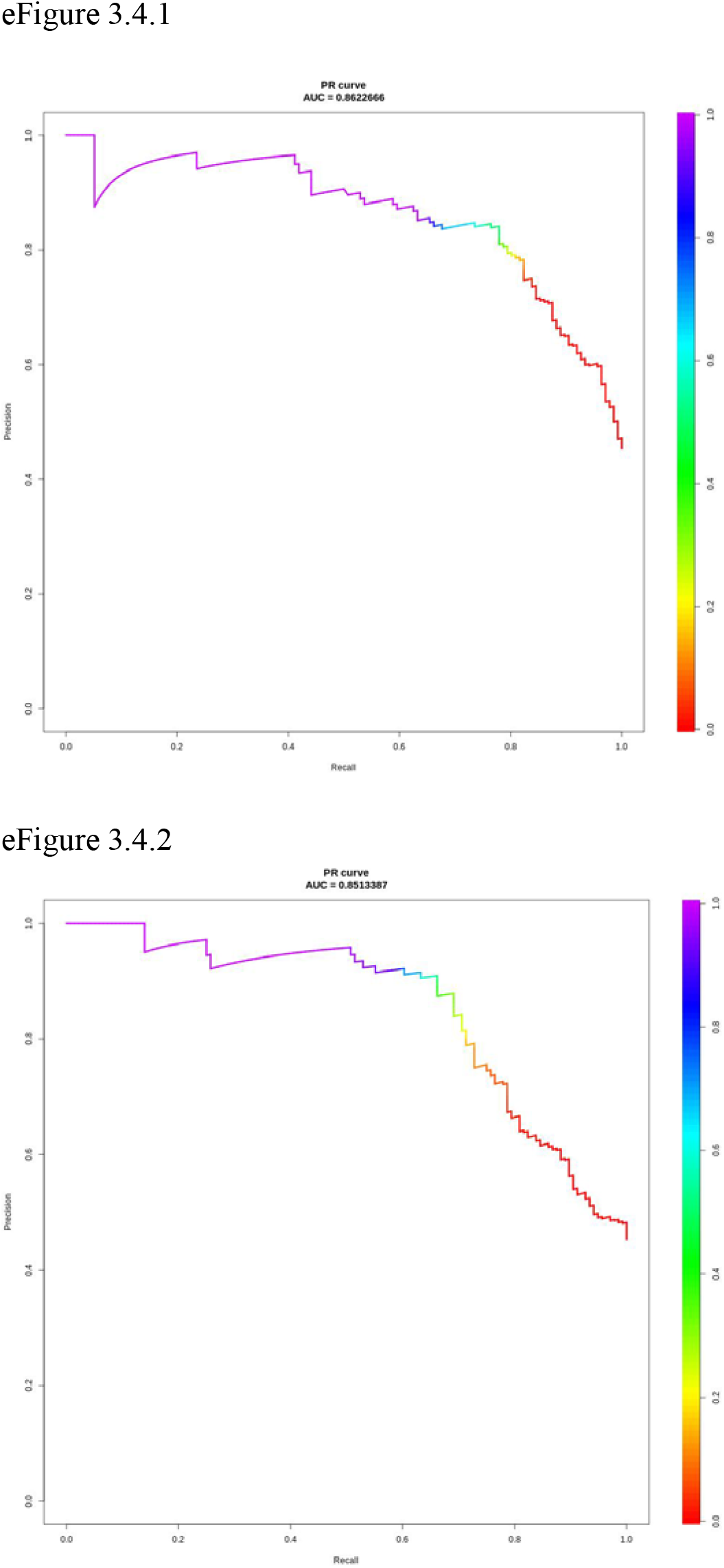
Precision-recall (PR) plots for the eight models. e2.1.1 – BERT-base, sliding window; e2.1.2 – BERT-base, hierarchical Transformers; e2.2.1 – ClinicalBERT, sliding window; e2.2.2 – ClinicalBERT, hierarchical Transformers; e2.3.1 – BERT-large, sliding window; e2.3.2 – BERT-large, hierarchical Transformers; e2.4.1 – Longformer, sliding window; e2.4.2 – Longformer, hierarchical Transformers. Color gradients indicate the decision threshold applied for label binarization, moving between a real value between 0 and 1.

